# Sex-specific associations of social determinants of health and genetic risk factors with atherosclerotic cardiovascular diseases incidence in the general population

**DOI:** 10.1101/2025.08.06.25333117

**Authors:** Audrey Paulin, Louis-Jacques Ruel, Sébastien Thériault, Benoit J. Arsenault

## Abstract

**Background and aims:** The combined contribution of polysocial risk factors (socioeconomic status, psychosocial factors and living environment) and genetic background on atherosclerotic cardiovascular disease (ASCVD) risk remains unknown. We investigated the contribution of a comprehensive polysocial risk score (PsRS) and polygenic risk scores (PRS) on ASCVD incidence.

**Methods:** We developed a PsRS using latent class analysis based on socioeconomic factors, psychosocial factors and living environment in 321,016 UK Biobank participants free of ASCVD. Participants were divided into three PsRS groups. The impact of the PsRS on incident ASCVD was assessed using Cox proportional hazards. The impact of the PsRS and coronary artery disease (CAD)PRS and ischemic stroke (IS)PRS on the incidence of CAD and IS, respectively, were also assessed.

**Results:** During a median follow-up of 12.5 years, 17,737 ASCVD events were recorded. Compared to participants with a low PsRS, those with a high PsRS had a higher risk of ASCVD (HR=1.46 [95% CI, 1.40-1.52], *p*<0.001). Risk associated with an elevated PsRS was higher for females compared to males. Compared to participants with a low PsRS in the bottom tertile of CAD PRS, those with a high PsRS in the top tertile of CAD PRS were at higher CAD risk (HR=4.24 [95% CI, 3.94-4.55], *p*<0.001). Similar results were obtained for IS.

**Conclusions:** A comprehensive PsRS was associated with incident ASCVD, particularly in females, and may exacerbate genetic susceptibility to both CAD and IS, suggesting that addressing polysocial risk factors is key to implementing preventive ASCVD strategies in the general population.

## Introduction

Atherosclerotic cardiovascular diseases (ASCVD) such as coronary artery disease (CAD) and ischemic stroke (IS) are amongst the leading causes of mortality worldwide.^1,2^ Many studies have underscored the impact of lifestyle and cardiometabolic health on the incidence of ASCVD and how modification of these risk factors can decrease long-term risk of ASCVD.^3–7^ In addition to traditional risk factors, genetic risk factors also play a critical role in the development of atherosclerosis and long-term ASCVD risk.^8,9^ Combined, these factors may play an increasingly important role in the long-term risk of ASCVD.

In addition to clinical, lifestyle and genetic risk factors, social determinants of health (SDOH) have been linked with ASCVD incidence and are increasingly recognized as important contributors to global cardiovascular health.^10^ There are multiple facets of SDOH that seem to influence ASCVD risk such as social and community, education, economic stability, neighborhoods and environment as well as healthcare access. Despite knowledge on the importance of including SDOH in cardiovascular research, most studies still define SDOH as the socioeconomic status alone instead of all aspects of SDOH. According to the Centers for Disease Control, he five SDOH domains include economic stability, education access and quality, health care access and quality, neighborhood and built environment, and social and community context.^11^ These represent key societal factors that influence health outcomes across the lifespan such as ASCVD.

A recent American Heart Association statement underscored the importance of including SDOH in cardiovascular research, especially since these factors may lie upstream of lifestyle-related risk factors for ASCVD.^12^ Recent finding indicates that SDOH may modulate genetic susceptibility to disease, reinforcing the necessity of integrating social factors in biomedical research.^13^ However, the joint impact of SDOH and genetic risk factors on the incidence of ASCVD remains underexplored, particularly in women.

The aim of this study is to investigate the relationship between a comprehensive set of SDOH, lifestyle-related risk factors and the incidence of ASCVD in females and in males and to investigate the joint association of SDOH and genetic risk factors on CAD and IS risk in participants of the UK Biobank free of ASCVD at baseline.

## Methods

### Study population

The UK Biobank is a large prospective cohort study including over 500,000 participants aged between 40 to 69 years old.^14^ All participants were recruited between 2006 and 2010 and provided written consent at the baseline assessment at one of 22 assessment centers across the United Kingdom. This resource provides researchers information on lifestyle, health status and genetics through questionnaires, interviews, physical measures, and blood and urine samples. The UK Biobank received approval from the British National Health Service, Northwest-Haydock Research Ethics Committee (16/NW/0274). Data access permission for this study was granted under UK Biobank application 25205. The study sample included 321,016 participants excluding participants with missing data on lifestyle and metabolic markers, a C-reactive protein (CRP) level ≥20 mg/L, as participants with CRP levels ≥20 mg/L might be more likely to have acute rather than systemic inflammation. Another analysis was performed in 318,022 participants with available genetic information. Participants who had a ASCVD event before recruitment were excluded from the study.

### Assessment of social determinants of health

To comprehensively assess SDOH, we developed a polysocial risk score (PsRS) based on socioeconomic factors, psychosocial factors and living environment as previously described.^15,16^ The socioeconomic exposure included total household income before tax (data-field 738), education level (data-field 6138), an education score (England: data-field 26414, Scotland: data-field 26431 and Wales: data-field 26421) and current employment status (data-field 6142). Psychosocial factors included the number of people in the household (data-field 709), social support (data-field 2110), number of social activities (data-field 6160), frequency of friends/family visits (data-field 1031), emotional distress (data-field 6145) and diagnosed psychiatric disorder (data-field 20002, 20544, 20126, 20480, 20485). The living environment part of the PsRS was defined using the Townsend deprivation index (data-field 189), a crime score (England: data-field 26416, Scotland: data-field 26434, Wales: data-field 26425), a housing score (England: data-field 26415, Scotland: data-field 26432, Wales: data-field 26423), type of accommodation (own or rent)(data-field 680), proximity of green space (data-field 24500), the proximity of blue space (data-field 24502) and the proximity of natural environment (data-field 24506). Social determinants of health information were obtained from the Touchscreen questionnaire at baseline. Participants were then divided in three groups using latent class analyses (LCA) using R package “poLCA”.^17^ LCA is a statistical procedure used to identify subgroups in a population based on different variables as SDOH.^18^ LCA have been previously explored for the analysis of socioeconomic status.^16^

### Cardiovascular health score

We developed a cardiovascular health score (CVHS) as previously described.^19^ The CVHS is based on four lifestyle factors and six cardiometabolic risk factors. Lifestyle parameters used in the CVHS included smoking status, fruits and vegetables intake, physical activity level and sleep quality. The cardiometabolic risk factors included in the CVHS included systolic blood pressure (SBP), diastolic blood pressure (DBP), CRP levels, triglyceride levels, LDL-cholesterol levels, and glycated hemoglobin (HbA_1C_). Body mass index (BMI) was added to the score because of its inclusion in several risk scores, including the Life’s Essential 8.^6^ One point was assigned for each recommendation achieved, for a maximal score of 11 and a minimal score of 0. Participants were than categorized in four groups, 9-11 as healthy, 7-8 as moderately, 5-6 as moderately unhealthy and 0-4 as unhealthy.

### Coronary artery disease and ischemic polygenic risk scores

We used CAD and IS polygenic risk scores (PRS) to evaluate the contribution of genetic factors on ASCVD incidence across PsRS groups. For this study, we used the CAD-PRS and the IS-PRS provided by the UK Biobank.^20,21^ We used these outcomes separately as no combined PRS is available in the UK Biobank.

### Study outcomes

The study outcome was incident ASCVD defined as first occurrences or death with the International Classification of Diseases, 10^th^ revision (ICD-10) codes for ischemic stroke (IS; I63) and myocardial infarction (MI; I21-I23). We also added to the ASCVD definition first surgical procedures with Office of Population, Censuses and Surveys: Classification of Interventions and Procedures, version 4 (OPSC-4) for coronary artery bypass grafting (K40.1-40.4, K41.1-41.4, K45.1-45.5), for coronary angioplasty, with or without stenting (K49.1-49.2, K49.8-49.9, K50.2, K75.1-75.4, K75.8-75.9). In genetic analyses, the CAD and IS component of ASCVD were studied separately. CAD includes MI, coronary artery bypass grafting and coronary angioplasty. Dates and causes of hospitalization or death were obtained from death registry, primary care information and hospital admission medical reports.

### Statistical analyses

Multivariable Cox proportional hazard models were used to evaluate the impact of PsRS on ASCVD incidence. We also used multivariable Cox proportional hazard models to establish the impact of a CAD-PRS on the incidence of CAD across PsRS categories as well as the impact of an IS-PRS on the incidence of IS across PsRS categories. Total and sex-specific hazard ratios (HRs) with their 95% confidence intervals (95% CI) were obtained from those analyses. All analyses were adjusted for age, self-report ethnicity and sex (in analysis including females and males). Dates of recruitment were used as the starting time and participants with event before recruitment were excluded. The end of the follow-up was the date of occurrence of the first event (ASCVD), death or censoring. If participants had no event during the follow-up and were still alive, the end of the follow-up was the last day of October 2021. The median follow-up was 12.5 years (interquartile range: 11.7-13.2). Schoenfeld tests were performed as a prior to Cox regressions to verify proportionality assumption and Schoenfeld residuals were visually inspected. All analyses were performed for all participants, females and males separately. All statistical analyses were conducted using R (v4.1.3).

## Results

### Baseline characteristics of study participants

The baseline characteristics of UK Biobank participants across PsRS groups defined by LCA are presented in Table 1. The mean age was 56.5 and 44% of participants were males. Of the 321,016 participants included in this study, 33.7% had a low PsRS, 46.5% had an intermediate PsRS and 19.8% had a high PsRS. Compared to participants in the low PsRS group, participants in the high PsRS group tend to be slightly older, have higher chances of being an active smoker, consume less fruits/vegetables and have lower physical activity levels. A higher proportion of participants in the high PsRS used antihypertensive medications (26.8%) and cholesterol-lowering medications (22.2%) compared to participants in the low PsRS group (15.8% and 12.5% respectively).

**Table 1.** Baseline characteristics of the UK Biobank participants selected for this study by polysocial risk score (PsRS).

| **Factor** | **Total** | **Low PsRS** | **Intermediate PsRS** | **High PsRS** |
| --- | --- | --- | --- | --- |
| N | 321016 | 108147 | 149370 | 63499 |
| Male* | 141116 (44.0%) | 47974 (44.4%) | 66201 (44.3%) | 26941 (42.4%) |
| Age, years, mean (SD) | 56.5 (8.1) | 54.9 (8.1) | 56.8 (7.9) | 58.4 (7.9) |
| White British* | 306469 (95.5%) | 101245 (93.6%) | 146774 (98.3%) | 58450 (92.0%) |
| BMI, kg/m<sup>2</sup>, mean (SD) | 27.3 (4.7) | 26.9 (4.6) | 27.1 (4.4) | 28.5 (5.4) |
| Current smoking* | 31724 (9.9%) | 9589 (8.9%) | 10406 (7.0%) | 11729 (18.5%) |
| Low fruit and vegetable intake* | 59104 (18.4%) | 18665 (17.3%) | 25243 (16.9%) | 15196 (23.9%) |
| Low physical activity* | 138785 (43.2%) | 45891 (42.4%) | 62163 (41.6%) | 30731 (48.4%) |
| Poor sleep quality* | 134083 (41.8%) | 42556 (39.4%) | 58849 (39.4%) | 32678 (51.5%) |
| Systolic BP, mm Hg, mean (SD) | 139.9 (19.7) | 137.6 (19.3) | 140.7 (19.6) | 141.9 (20.1) |
| Diastolic BP, mm Hg, mean (SD) | 82.4 (10.7) | 81.9 (10.7) | 82.6 (10.6) | 82.8 (10.8) |
| Antihypertensive medication* | 61225 (19.1%) | 17077 (15.8%) | 27116 (18.2%) | 17032 (26.8%) |
| CRP, mg/L, mean (SD) | 2.6 (4.3) | 2.3 (4) | 2.4 (4.1) | 3.4 (5.1) |
| Triglycerides, mmol/L, mean (SD) | 1.7 (1.0) | 1.7 (1) | 1.7 (1) | 1.9 (1.1) |
| LDL-C, mmol/L, mean (SD) | 3.6 (0.9) | 3.6 (0.8) | 3.6 (0.9) | 3.5 (0.9) |
| HDL-C, mmol/L, mean (SD) | 1.5 (0.4) | 1.5 (0.4) | 1.5 (0.4) | 1.4 (0.4) |
| Cholesterol lowering medication* | 48366 (15.1%) | 13476 (12.5%) | 20763 (13.9%) | 14127 (22.2%) |
| HbA1c, mmol/mol, mean (SD) | 35.9 (6.5) | 35.4 (5.9) | 35.6 (5.9) | 37.5 (8.1) |

\* n (%)

### Cross-sectional associations of social determinants of health and clinical and lifestyle-related risk factors

To investigate the relationship between SDOH and clinical and lifestyle-related risk factors at baseline, we investigated the distribution of the CVHS across the three PsRS categories (Figure 1). Among participants classified as having a healthy CVHS, 39% had a low PsRS, and only 12% a high PsRS. In contrast, in participants with an unhealthy CVHS, only 25% were characterized by a low PsRS, whereas 39% with a high PsRS. These results indicate that individuals with a high PsRS are less likely to have optimal management of cardiometabolic risk factors and less likely to have a healthy lifestyle.

**Figure 1.**
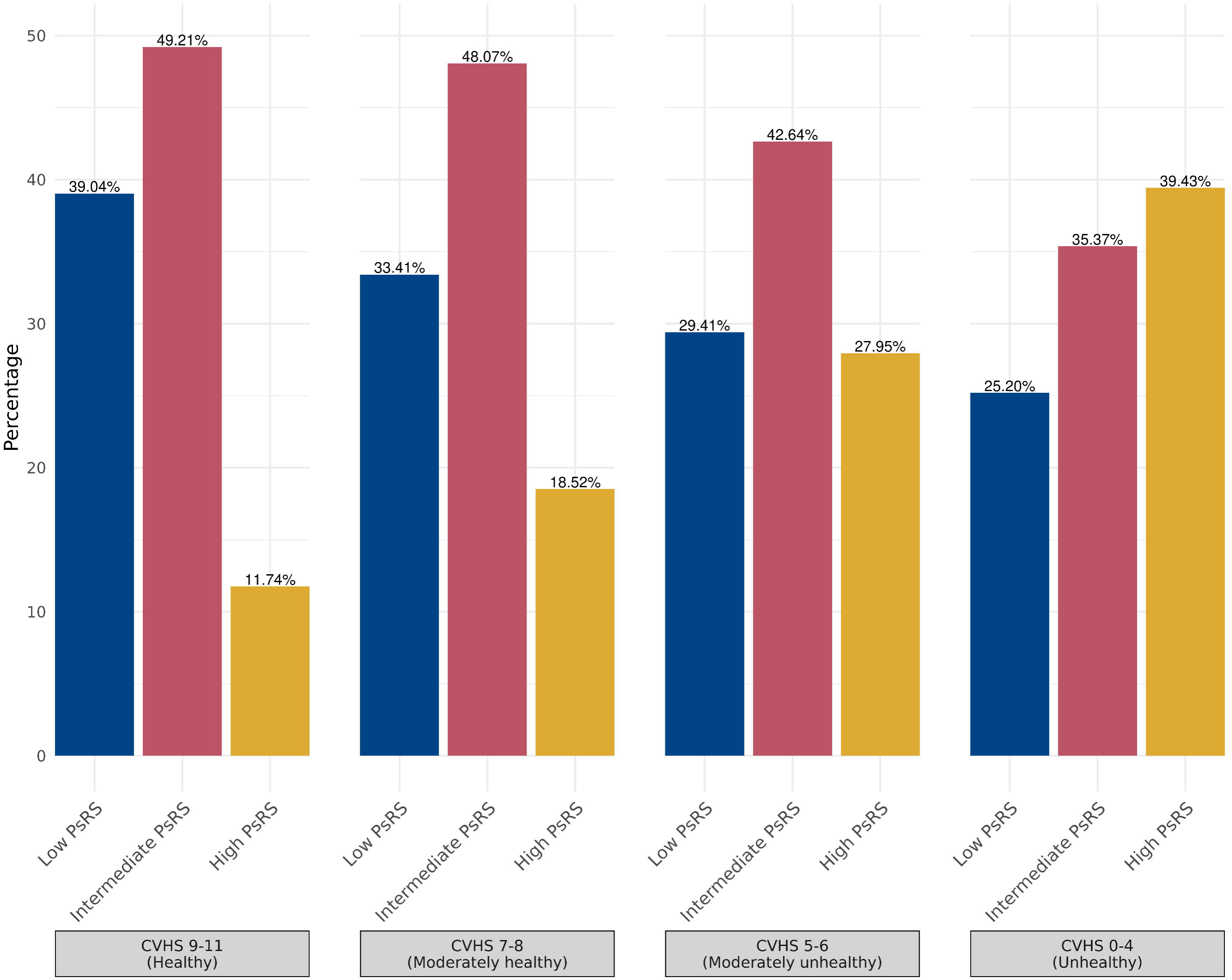
Distribution of UK Biobank participants in each polysocial risk score (PsRS) categories across the cardiovascular health score (CVHS).

### Sex-specific impact of the polysocial risk score on ASCVD incidence

During the study follow-up, 17,737 study participants had a cardiovascular event (defined in methods). We investigated the impact of the PsRS on the incidence of ASCVD before and after adjusting for clinical and lifestyle-related risk factors using multivariate Cox regression models (Figure 2). The model 1 included the PsRS, age, sex and ethnicity while the model 2 included the PsRS, age, sex, ethnicity and components of the CVHS. For the model 1, compared to participants with a low PsRS risk, those with an intermediate PsRS risk had a slightly higher ASCVD risk (hazard ratio [HR] = 1.09 [95% CI, 1.02-1.16], *p*=0.009) while those with a high PsRS risk had a 78% increase ASCVD risk (HR = 1.78 [95% CI, 1.66-1.90], *p*<0.001). For the model 2, compared to participants with a low PsRS risk, those with an intermediate PsRS risk had a nonsignificant slightly higher ASCVD risk (HR = 1.04 [95% CI, 0.97-1.11], *p*=0.24) while those with a high PsRS risk still had a higher ASCVD risk (HR = 1.38 [95% CI, 1.29-1.47], *p*<0.001). Although trends were similar in females versus males, the effect of the PsRS on ASCVD incidence was more pronounced in females (HR=1.69 [95% CI, 1.58-1.81], *p*<0.001 and HR=1.46 [95% IC, 1.39-1.54], *p*<0.001, respectively, for females and males with a high PsRS compared to participants with a low PsRS within the same sex with an interaction p-value = 3.65e-07). Altogether, these results suggest that individuals with a high PsRS are at higher risk of ASCVD, that this association was stronger in females compared to males and that clinical and lifestyle-related risk factors had a modest effect on the relationship between higher PsRS and ASCVD risk.

**Figure 2.**
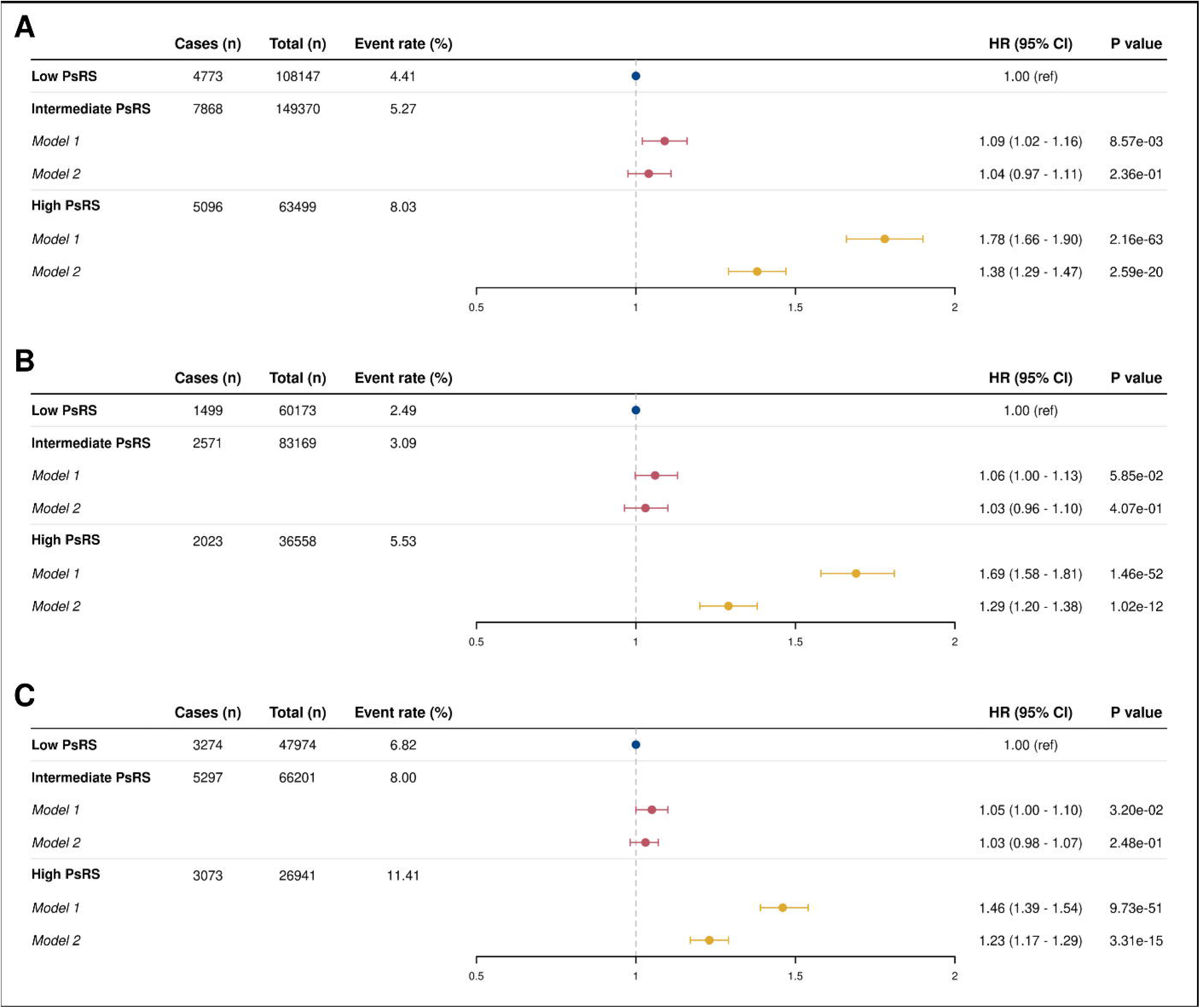
Impact of a polysocial risk score (PsRS) on the incidence of atherosclerotic cardiovascular diseases. Results are presented in: A) all participants, B) females and C) males. Model 1 is adjusted for age, sex, and ethnicity while model 2 is adjusted for age, sex, ethnicity, and components of a cardiovascular health score.

### Sex-specific joint associations of the polysocial risk score and a coronary artery disease polygenic risk score on coronary artery disease incidence

During the study follow-up, 12,861 study participants developed coronary artery disease. Figure 3 presents the impact of a polygenic risk score for coronary artery disease (CAD-PRS) on the incidence of CAD in each PsRS categories. Overall, a graded increase in CAD risk was observed across CAD-PRS tertiles in each PsRS categories. When compared to individuals with a low PsRS in the bottom CAD-PRS tertile, the CAD risk in participants with a low PsRS in the top CAD-PRS tertile (HR = 3.05 [95% CI: 2.78– 3.33], *p*<0.001) was higher than those with a high PsRS in the bottom CAD-PRS tertile (HR = 1.64 [95% CI: 1.47–1.82], *p*<0.001), suggesting a additive effect of both genetic and SDOH. Within each PsRS groups, an increase CAD risk was observed with increasing CAD-PRS, with the highest risk noted in individuals in the top tertile of CAD-PRS and high PsRS (HR = 4.36 [95% CI: 3.99–4.77], *p*<0.001). Comparable results were observed in both females and males (Figure 3b and 3c). These results highlight that genetic risk factors for CAD predict risk independently of SDOH and that the highest risk of CAD is observed in patient with high polysocial and genetic risk.

**Figure 3.**
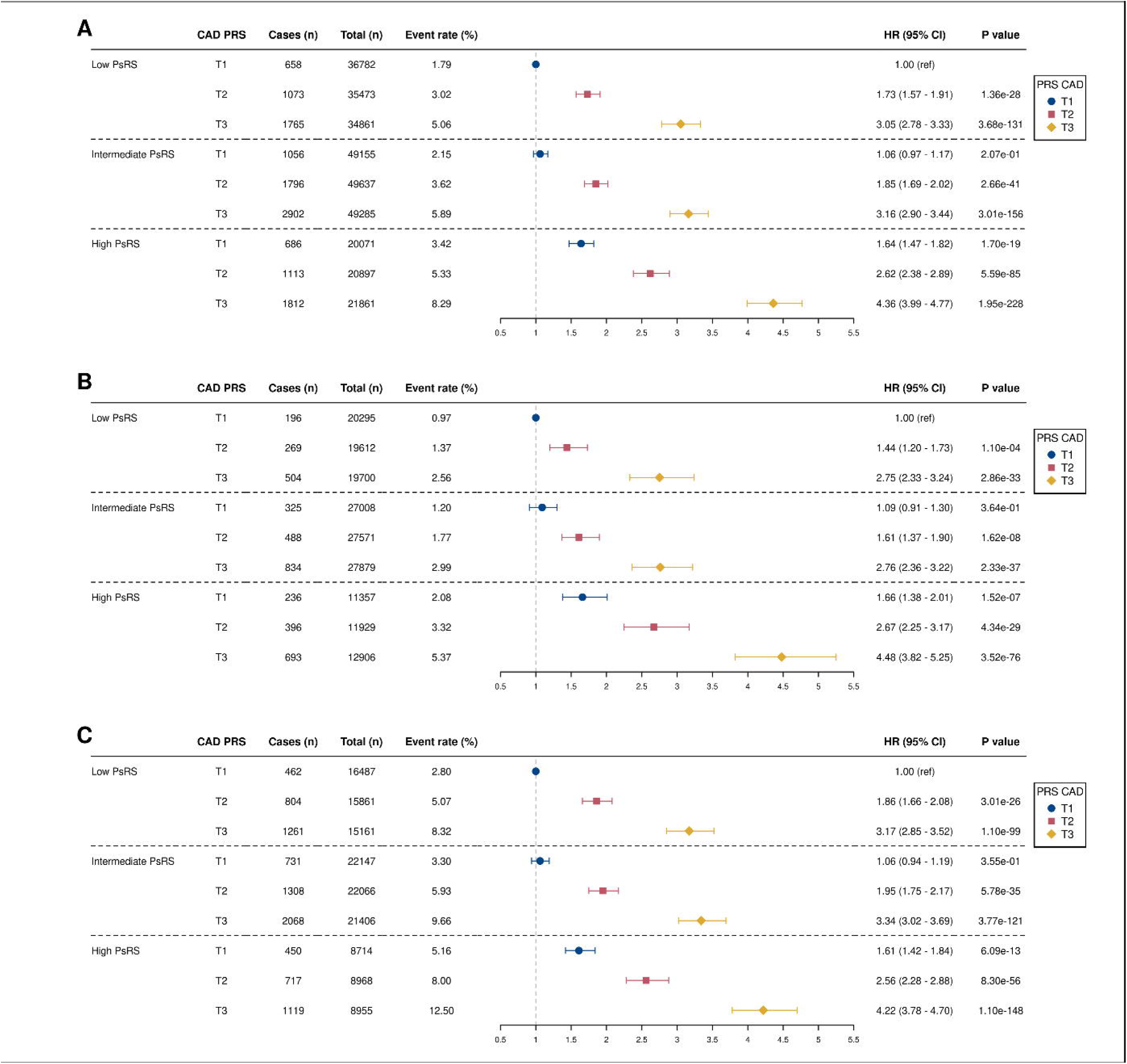
Joint contributions of a polysocial risk score (PsRS) and a polygenic risk score for coronary artery disease (CAD-PRS) on the incidence of CAD. Results are presented in: A) all participants, B) females, and C) males. Cox regression analyses are adjusted for age, sex, and ethnicity.

### Sex-specific joint associations of the polysocial risk score and an ischemic stroke polygenic risk score on ischemic stroke incidence

During the study follow-up, 5104 study participants had an ischemic stroke event. Figure 4 presents the impact of an IS-PRS on the incidence of IS in each PsRS category. Within each PsRS group, increasing IS-PRS tertiles were associated with a higher risk of IS. Participants with a high PsRS in the lowest IS-PRS tertile (HR = 1.72 [95% CI: 1.48– 1.99], *p*<0.001) had a similar risk to those with a low PsRS in the top IS-PRS tertile (HR = 1.92 [95% CI: 1.67–2.20], p < 0.001). Compared to participants with a low PsRS in the bottom IS-PRS tertile, those with a higher PsRS in the top tertile of IS-PRS had the highest IS risk (HR = 2.76 [95% CI: 2.42–3.15], *p*<0.001). Comparable results were observed in both females and males (Figure 4b and 4c). Altogether, these results suggest that SDOH may be as important as genetic factors to predict IS risk and that the combination of SDOH and genetic factors significantly increase IS risk.

**Figure 4.**
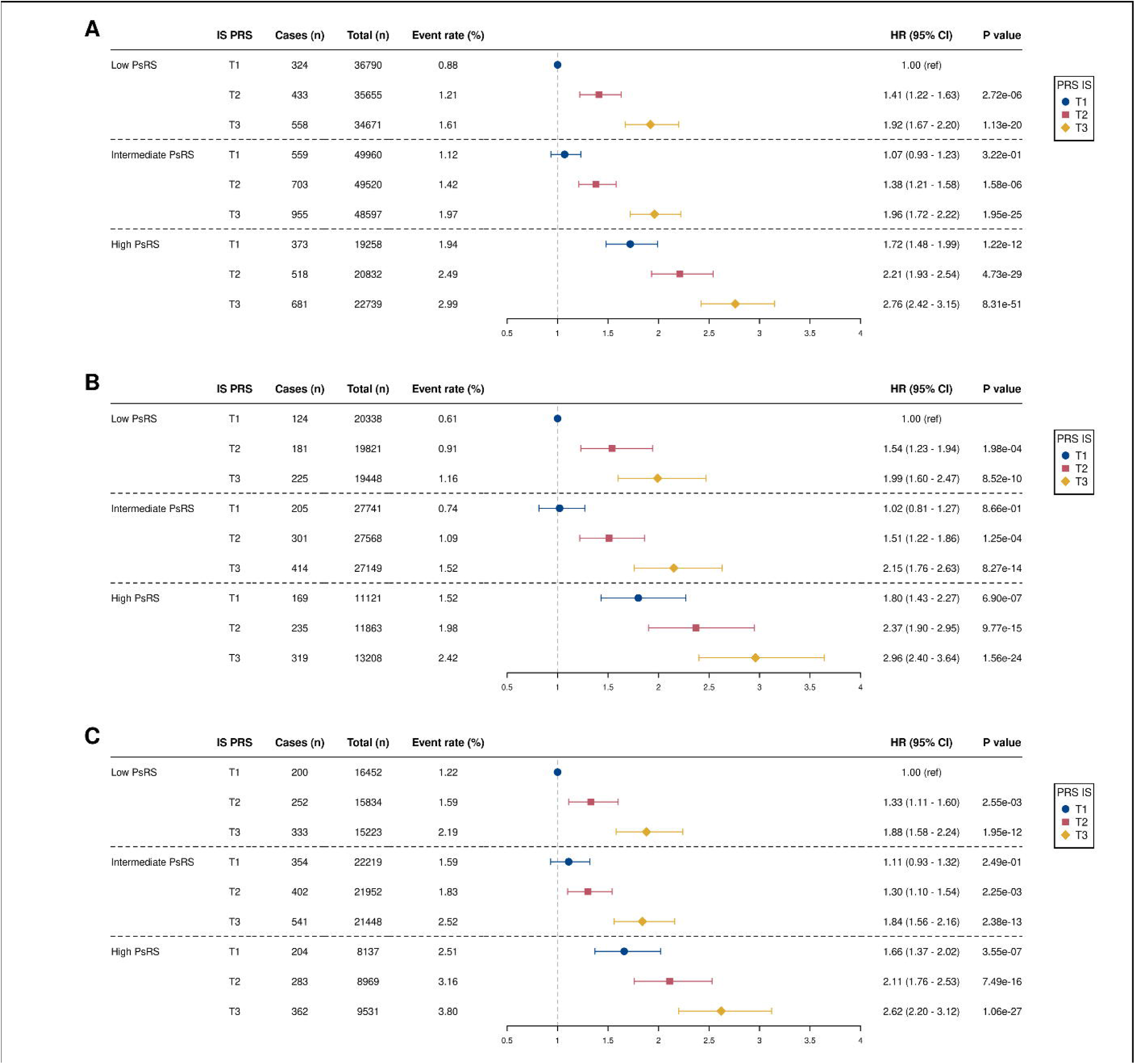
Joint contributions of a polysocial risk score (PsRS) and a polygenic risk score for ischemic stroke (IS-PRS) on the incidence of IS. Results are presented in: A) all participants, B) females, and C) males. Cox regression analyses are adjusted for age, sex, and ethnicity.

## Discussion

To better understand the role of multiple SDOH in the prediction of ASCVD, we developed a comprehensive PsRS that considered socioeconomic factors, psychosocial factors such as wellbeing and factors related to the living environment. We tested the impact of this PsRS on ASCVD events in a large sample of participants of the UK Biobank. Our results suggest that individuals with a high PsRS are three times less likely to have a healthy CVHS than those with a low polysocial risk. The PsRS was associated with ASCVD incidence, even when lifestyle and metabolic health-related risk factors were considered. Females in the high PsRS group had a higher risk of ASCVD compared to males of the same group. Finally, SDOH also modulate the impact of genetic susceptibility on ASCVD risk, the latter being especially relevant to IS as SDOH (PsRS) appeared to have an effect on IS risk comparable to genetic risk factors (IS-PRS).

Our results underscore the importance of not only including SDOH when assessing individual risk of ASCVD but also addressing SDOH in primordial prevention of ASCVD to promote the adoption of healthier lifestyle habits and optimal management of ASCVD risk factors. These results support that SDOH have additional effect on traditional risk factors for the prevention of ASCVD. Different research groups focused on the effect of adding SDOH to existing scores such as the Framingham Risk Score.

Results of those studies suggest that SDOH improved the stratification of high-risk individuals.^22–25^ Our results are also consistent with other studies showing that individuals living in a more favorable environment tend to have lower alcohol intake and smoking, and higher physical activity levels, sleep quality and diet quality.^15,22,26,27^ Interventions targeting individual behaviors and metabolic risk factors, while essential, may be insufficient without broader structural efforts to mitigate risk associated with SDOH. Policies aimed at reducing income inequality, improving access to education, and ensuring stable housing can help address the foundational socioeconomic disparities that contribute to elevated ASCVD risk.^10,22,28,29^ Psychosocial factors such as chronic stress, social isolation, and low perceived well-being also warrant targeted interventions, including community-based programs that promote social cohesion, mental health services, and workplace wellness initiatives. Environmental improvements, such as creating safe, walkable neighborhoods and increasing access to healthy food options, are equally critical.

In our study, females in the high PsRS group had a higher risk of ASCVD than males in the high PsRS group when compared to individuals in the low PsRS group. These results are consistent with the work of Yang et al. In this study of 40,536 participants from the National Health and Nutrition Examination Survey (NHANES), females with the highest level of cumulative unfavorable SDOH had a sixfold increased risk of premature ASCVD compared to females without unfavorable SDOH. Men had a threefold increased risk of premature ASCVD for the same group comparation.^30^ A meta-analysis of over 22 million individuals (not including participants of the UK Biobank) also reported similar results on sex difference when studying the relationship between socio-economic status and cardiovascular diseases.^31^ The stronger association between SDOH and ASCVD among women observed in our study may reflect both biological and social mechanisms.

Women are disproportionately affected by adverse social conditions such as lower income, single parenthood, and caregiving responsibilities, which can compound psychosocial stress and limit access to health-promoting resources.^32,33^ Chronic stress and social adversity have been shown to impact cardiovascular health through neuroendocrine and inflammatory pathways, potentially interacting with hormonal and vascular differences unique to females.^34^ Moreover, traditional cardiovascular risk models may underrepresent the cumulative burden of psychosocial stressors more common in women. Structural and gender-based inequities may also lead to delays in diagnosis and treatment, amplifying the effects of social disadvantage.

Finally, our findings also reveal that SDOH may modulate the impact of genetic susceptibility on ASCVD, particularly for IS. The observation that the effect of polysocial risk was comparable in magnitude to that of the IS-PRS suggests that unfavorable social conditions can amplify or potentially offset inherited genetic risk. This highlights the importance of considering both genetic and social risk factors in ASCVD prevention strategies. From a precision medicine perspective, integrating SDOH into risk stratification models may improve prediction and help identify individuals who would benefit most from early intervention, especially those with high genetic risk who are also socially disadvantaged. Moreover, our results support the growing recognition that genetic risk is not deterministic and can be influenced by modifiable environmental and social contexts.^9,35^ These insights reinforce the need for multidisciplinary approaches that bridge genomics with public health and social policy.

Strengths of the current study include the large cohort of individuals aged between 40 to 69 years old. This cohort offers a large variety of SDOH allowing the inclusion of the five domains of SDOH. We also used a very comprehensive PsRS as SDOH have a cumulative impact on the incidence of ASCVD.^29^ This study also has several limitations. The study population was predominantly White, slightly healthier than the general population and lived in less socioeconomically deprived areas.^36^ In our study population, only 19.8% participants had a high PsRS. Also, we only investigated the impact of the PsRS on the incidence of ASCVD. The PsRS could also influence other chronic diseases such as type 2 diabetes incidence.^15,37^ Finally, this prospective study was observational by design and causality cannot be inferred. Bias due to unmeasured confounders and reverse causality cannot be excluded.

In conclusion, results of the current investigation highlight the need for multifactorial and intersectoral approaches targeting SODH to improve the adoption of healthier lifestyles. and management of clinical risk factors to reduce the burden of ASCVD in the general population. Addressing SDOH represents a promising, equity-oriented path toward reducing the burden of ASCVD particularly in women and in genetically susceptible individuals.

## Declarations

### Institutional Review Board Approval

All participants provided written consent at the baseline assessment at one of 22 assessment centers across the United Kingdom. The UK Biobank received approval from the British National Health Service, Northwest-Haydock Research Ethics Committee (16/NW/0274).

### Data Availability

Access to UK Biobank data can be granted via the Access Management System of the UK Biobank (https://www.ukbiobank.ac.uk/enable-your-research/apply-for-access).

### Code Availability

The code used to perform this analysis will be released on GitHub upon acceptance of the manuscript.

### Competing interests

BJA is a consultant for Novartis, Eli Lilly, and Silence Therapeutics and has received research contracts from Pfizer, Eli Lilly and Silence Therapeutics.

### Funding

The UK Biobank was established by the Wellcome Trust medical charity, Medical Research Council, Department of Health, Scottish Government and the Northwest Regional Development Agency. It has also had funding from the Welsh Government, British Heart Foundation, Cancer Research UK and Diabetes UK. HDM holds a doctoral research award from the Quebec Heart and Lung Institute. LJR is supported by a doctoral research award from the Canadian Institutes of Health Research. BJA holds a senior scholar award from the *Fonds de recherche du Québec*.

### Authors’ contributions

Data acquisition and analysis AP and ST. Conception and design AP, BJA. Drafting of the work AP, LJR and BJA. All authors approved the final version of the manuscript.

---

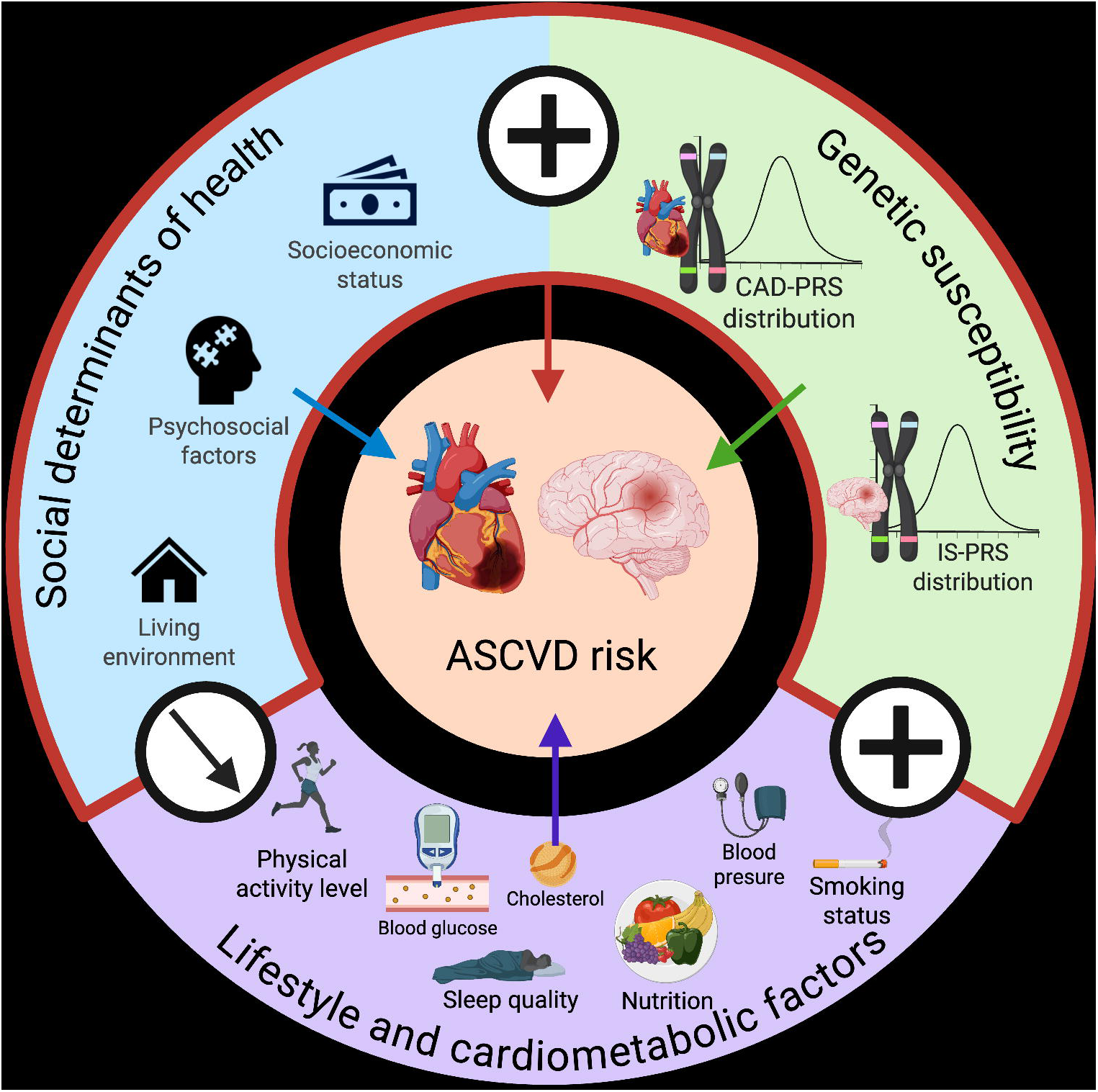

## Notes

### Author Declarations

All participants provided written consent at the baseline assessment at one of 22 assessment centers across the United Kingdom. The UK Biobank received approval from the British National Health Service, Northwest-Haydock Research Ethics Committee (16/NW/0274). Data access permission for this study was granted under UK Biobank application 25205.

